# Accurate and cost-efficient whole genome sequencing of hepatitis B virus using Nanopore

**DOI:** 10.1101/2024.08.12.24311345

**Authors:** Joakim B. Stenbäck, Daniel Schmidt, Ulrika Noborg, Joel Gustafsson, Peter Norberg, Maria E. Andersson, Michael X Fu, Heli Harvala, Johan Ringlander

**Affiliations:** Department of Infectious Diseases, Institute of Biomedicine, University of Gothenburg, Sweden; Department of Clinical Microbiolgy, Sahlgrenska University Hospital, Gothenburg, Sweden; Bioinformatics and Data Centre, University of Gothenburg, Sweden; Nuffield Department of Medicine, University of Oxford, United Kingdom; Radcliffe Department of Medicine, University of Oxford, United Kingdom

## Abstract

Deep sequencing of the whole hepatitis B virus genome increases the analytical resolution and has the potential to improve molecular epidemiology investigations. The aim of this work was to develop and evaluate the performance of such deep sequencing using the Nanopore technology. The method includes an initial PCR step to generate two overlapping amplicons that cover the whole relaxed circular HBV genome found in circulating viral particles and covalently closed circular DNA in infected hepatocytes, followed by sequencing using the Nanopore rapid barcoding kit that allows parallel analysis of several samples in one reaction. The libraries can be sequenced with the standard Nanopore flow cell on MiniIon or GridIon devices, as well as the Flongle. The performance of the method was evaluated by comparing Nanopore and Sanger sequences or qPCR results from 64 clinical samples. The Nanopore-derived consensus sequences were, on average, 99.9% similar to those from Sanger sequencing and the full HBV genome was determined in samples with HBV DNA levels of approximately 3 log_10_ IU/mL with MagNA pure 96 extraction and < 2 log_10_ IU/mL using a high-volume manual extraction protocol on a subset of samples from patients with very low viral load (1.62-3.74 IU/mL). A perfect agreement with Sanger/qPCR-derived genotype was seen. The cost of sequencing per genome using the Nanopore method is low, ranging 6-37euros. We conclude that whole-genome sequencing of HBV with Nanopore is well suited for genomic characterization, antiviral resistance mutation analysis and genotyping of HBV in a routine laboratory setting.

## Background

Chronic infection with hepatitis B virus (HBV) causes a heavy health burden worldwide (1). Chronic hepatitis B poses a risk for long-term complications such as cirrhosis and cancer, a risk that can be reduced by treatment with nucleot(s)ide analogs (2). HBV has nine genotypes (A-I) with relatively distinct geographical distribution, differing from each other by at least 8% of the ≈ 3200 nucleotides that comprise the genome. In addition, there are a large number of subgenotypes, some of which require whole genome comparison for accurate classification. Whole genome sequencing can also support molecular epidemiology investigations, for example, when evaluating potential transmission between subjects infected with the same genotype (3, 4). Identification of genotypes can be achieved by simplified methods such as real-time PCR (5), but new sequencing techniques have become almost as easy and allow relatively fast genotyping at a relatively low cost. Also, the short circular HBV genome enables full genome coverage by only two overlapping PCRs. Genotyping has clinical applicability since infection with genotype C is a risk factor for liver complications, and infection with genotype A is more susceptible to interferon treatment (6).

Although antiviral resistance against tenofovir and entecavir is relatively rare, resistance analysis is quite often of interest in patients with poor therapeutic response. Entecavir resistance typically occurs after previous treatment with lamivudine (7, 8). Resistance to tenofovir is very rare, but mutations leading to substitution 236T in reverse transcriptase, in combination with other mutations, have been suggested to lead to antiviral resistance (9, 10).

Nanopore sequencing has been widely used during the COVID-19 pandemic (11–14), and this technique is also increasingly used in bacteriology for antimicrobial resistance genotyping (15), 16s and whole-genome sequencing (16, 17). Many clinical microbiological laboratories that previously did not perform sequencing are now in possession of one or multiple Nanopore instruments (18). Apart from whole-genome sequencing of SARS-CoV-2 (19), methods for many other microbes have been launched (20–22). The relatively low upfront cost for the device, low cost of sequencing and simple protocols are benefits that have been described. On the other hand, a higher error rate, the need for high computer calculation capacity and specialized bioinformatics may discourage the introduction of the Nanopore method in a routine laboratory. In this article, we describe the development of a user-friendly method for Nanopore sequencing of HBV and a coherent bioinformatical pipeline adapted for clinical use in routine laboratories.

## Methods

### Patients and samples

Patient samples from three different cohorts were part of this study. Cohort 1 consisted of 38 HBV positive sera from individual patients that had been analyzed with a Sanger sequencing-based resistance testing method as part of routine diagnostics at the Department of Clinical Microbiology, Sahlgrenska University Hospital, Gothenburg. One sample from this cohort was quantified with qPCR and serially diluted to assess the analytical sensitivity. Cohort 2 was made out of 19 sera that had previously been genotyped using qPCR (5) in routine diagnostics, also at Sahlgrenska University Hospital. Cohort 3 consisted of 7 plasma samples from blood donor screening that all had low viral load and previously had been quantified using ultrasensitive PCR and genotyped using a Sanger sequencing-based method in the United Kingdom, as previously described (23). Details on the number of samples, genotype and viral load are found in Table 1.

**Table 1.** Number of patients, genotype and viral load among the study cohorts.

| **HBV genotype** | **Cohort 1** | **Cohort 2** | **Cohort 3** | **Total** |
| --- | --- | --- | --- | --- |
| A | 5 | 0 | 2 | 7 |
| B | 2 | 6 | 1 | 9 |
| C | 8 | 8 | 1 | 17 |
| D | 19 | 4 | 1 | 24 |
| E | 3 | 1 | 2 | 6 |
| F | 1 | 0 | 0 | 1 |
| All | 38 | 19 | 7 | 64 |
| **Viral load IU/mL log<sub>10</sub>** |  |  |  |  |
| <3 | 2 | 0 | 4 | 5 |
| 3-5 | 24 | 0 | 3 | 28 |
| >5 | 12 | 19 | 0 | 31 |
| **Same genotype as Sanger/qPCR** | 100% | 100% | 100% | 100% |
| **Antiviral resistance sensitivity** | 100% | NA | NA | 100% |
| **Viral load IU/mL log<sub>10</sub>, mean** | 4.5 | 7.7 | 2.7 | 4.7 |
| **Fraction of the HBV genome, mean** | 93.7% | 100% | 99.8% | 99.9% |
| **Samples with full genome coverage (N/Total N)** | 33/38 | 19/19 | 7/7 | 59/64 |

### Nucleic acid extraction and HBV DNA quantification

For cohorts 1 and 2, which had been previously analyzed in routine diagnostics at the Sahlgrenska University Hospital, Gothenburg, Sweden, total NA extraction from 200 uL of blood/plasma was performed using the MagNA pure 96 (Roche, Basel, Switzerland), according to the manufacturer’s instructions. For the low viral load panel (cohort 3), large-volume extraction from 5 mL of plasma was made using a modified protocol for the Roche High Volume NA extraction kit, as previously described (23). Four samples in cohort 3 were analyzed using both extraction methods for comparison. For cohorts 1 and 2, HBV DNA quantification was performed using the Cobas HBV test on the 6800 platform (Roche) according to the manufacturer’s instructions; for cohort 3, quantification was performed as previously described (23).

Serial dilutions of one sample with an original concentration of 5.34 log_10_ IU/mL were sequenced with Nanopore to further assess the sensitivity. The sample was diluted in H_2_0 to 1:10, 1:100, 1:1000 and 1:10000. Only the first sets of primers for the whole genome method was used and the nested PCR covering the RT region was not applied. The dilutions were sequenced on a flongle flow cell.

### PCR and Nanopore sequencing

PCRs intended to generate two overlapping amplicons covering the whole genome were designed. The primers used for amplicon 1 were 56F (CCTGCTGGTGGCTCCAGT) and 1827R (GAAAAAGTTGCATGGTGCTGGT), and for amplicon 2 primers 1689F (ACCGACCTTGAGGCCTACTTCA) and 262R (CCACCACGAGTCTAGACTCT). Primers were manually designed based on an alignment with whole genomes of genotypes A-G. Mismatches in primers targets were allowed and the sequences were chosen to preferably target A-E, the most common genotypes in Sweden and the UK. Identical settings and reagents were used in the two amplifications. To the reaction, 5 µL of sample and 1 µL of each primer in 10 µM dilutions were added to 10 µL of master mix containing Hot Start PCR mix (2X) (ThermoFisher, Waltham, MA, USA) and 10 µL H_2_O. A denaturation step at 95°C was included prior to 40 cycles of PCR, with annealing at 60°C for 10 s and elongation at 72°C for 2 min.

Capillary gel electrophoresis was performed on a TapeStation 4200 (using the DT5000 kit, Agilent, Santa Clara, CA, USA), and if no visible bands were seen, a nested PCR with inner primers 251F (GTGGTGGACTTCTCTCAATTTTC) and 1801R (CAGACCAATTTATGCCTACAGCCT) was performed with the same PCR settings as described in the first PCR. In this study, only the amplicon covering the reverse transcriptase (RT) region was nested.

Amplicons were barcoded, and libraries were prepared with the Rapid Barcoding Kit 96 SQK-RBK110.96, attaching barcodes and adapters randomly on the amplicons using transposase (both kits from Oxford Nanopore Technologies, Oxford, UK). Samples were pooled and sequenced with the MinIon sequencing device (Oxford Nanopore Technologies) and the R10.4.1 flowcell (FLO-MIN114) or an R10.4.1 flongle (FLO-FLG114) (Oxford Nanopore Technologies). Cohort 1 samples were sequenced five at a time on a flongle flowcell to mimic the planned use in routine diagnostics. Cohort 2 and 3 samples were sequenced on one standard flowcell each, and reruns for reproducibility testing of cohort 3 samples were performed using the flongle. One or two negative controls (H2O) were used in PCR and subsequent Nanopore runs and bioinformatic analysis.

This protocol is available at protocols.io (https://www.protocols.io/view/hbv-whole-genome-sequencing-using-oxford-nanopore-n92ldrow9g5b/v1).

### Sanger sequencing

Parts of the reverse transcriptase (RT) region was sequenced with the Sanger method. For cohort 1, Sanger sequencing was performed after PCR with primers 593F (TGCACCTGTATTCCCATCCCATC) and 1010R (GRGCAGCAAADCCCAAAAGACC), if no band was seen, semi-nested PCR was performed with 635F (TTCCTATGGGAGTGGGCCTC) and the same reverse. For cohort 3, parts of the RT region was covered by two amplicons and Sanger sequenced, as previously described (23).

### Bioinformatics

Base calling of Nanopore fast5 files was made with the GPU-accelerated guppy program (https://github.com/asadprodhan/GPU-accelerated-guppy-basecalling) with super accurate mode settings. Subsequently, fastq files were run through a pipeline including following steps: Reads were mapped to HBV reference genomes representing genotypes A-H using Minimap2. Draft consensus sequences were created using samtools and polished with Medaka (Oxford Nanopore Technologies) and the consensus sequence from the best matching genotype was chosen. Minor variant analysis was performed with freebayes (24) and a an inhouse script was used to call resistance mutations based on those listed at geno2pheno (https://hbv.geno2pheno.org/). Samples were assigned as passed or failed, the latter if fewer reads than the negative control. The pipeline is available at github (https://github.com/ClinicalGenomicsGBG/hbv_nano). The Stanford HBV database (https://hivdb.stanford.edu/HBV/HBVseq/development/hbvseq.pl?action=showSequenceForm) and geno2pheno tool (https://hbv.geno2pheno.org/) were used for genotyping and calling antiviral resistance mutations from consensus sequences and to compare sequence quality in the RT region, and showed consistent results with the pipeline. Splicing analysis was made using FLAME (25).

## Results

### Antiviral resistance analysis and genotyping

In cohort 1, 18/38 samples had previously detected antiviral resistance mutations by Sanger sequencing. The same mutations were found using Nanopore, with the addition of a mutation causing the resistance-associated amino acid substitution 80I in one sample, that only was found with the Nanopore method, as this position was not covered by the Sanger amplicon. Samples with resistance mutations had a viral load of 5.0 log_10_ IU/mL in mean (range 3-7.92 log_10_ IU/mL) All minor variants found with the Sanger-based analysis were also detected by and the antiviral resistance mutation with the lowest frequency was present in 25% of the Nanopore reads. All samples but four could be analyzed regarding antiviral resistance without performing nested PCR and those had low viral loads ranging 2.4-3.37 log_10_ IU/mL. Thus, the clinical sensitivity for antiviral resistance mutations was 100%. There were no antiviral resistance-associated mutations in cohorts 2 and 3, that previously not had been analyzed for resistance mutations.

In cohort 1,2 and 3, genotyping using Sanger or qPCR (cohort 2) did not differ from the Nanopore results. HBV genotypes A-F were represented among the samples (table 1). The number of samples with mismatches in the Nanopore method primer targets was low and typically only in one position. However, the four samples with an unsuccessful whole genome sequencing might have had more mismatches in primers 56F and 1827R, that could not be detected. Sequencing of the RT region of these samples after nested PCR was successful and showed a lack of mutations in 1689F and 262R primer targets for all but the one sample with genotype F that had multiple differences.

### Sensitivity and reproducibility

In cohort 1, the entire HBV genome was obtained in all samples >3.27 log10 IU/mL, that was 33/38 samples. In the one sample with genotype F, the whole amplicon covering the RT region, but not the whole genome, was obtained after the first PCR, thus, nested PCR was not performed. The remaining four samples (all ≤3.27 log10 IU/mL with genotype D (n=3) and genotype B (n=1)) were subject to nested PCR targeting the region spanning nt 251-1801(including the RT), and the entire region was obtained in all samples.

A sample with an original concentration of 5.34 log_10_ IU/mL was diluted and sequenced. Sequences were identical in dilutions down to 1:100 but the number of reads in 1:1000 and 1:10000 were insufficient for antiviral resistance analysis and assembly of a whole genome. At dilution 1:100, the full genome was not captured but the whole RT region was, which was identical to the RT region of the 1:1 and 1:10 dilutions. Thus, the analytical sensitivity of the method, using the standard MagNA pure 96 extraction and when five samples were pooled and run on one flongle flow cell, was approximately 3.34 log_10_ IU/mL. Table 2 shows results from the sequencing of a serially diluted sample. In cohort 2 all samples were successfully whole genome sequenced. For cohort 3, consisting of seven samples, of which four samples were extracted with both the automated MagNA pure 96 protocol and the large-volume manual extraction protocol, the full HBV genome was successfully Nanopore sequenced after manual extraction in all samples. The full genome was captured in 3/4 of the samples after MagNA pure 96 extraction, and in the fourth sample, only the amplicon covering the RT region showed reads. The three successful samples had viral loads of 3.43, 3.27 and 1.74 log_10_ IU/mL, respectively, and the remaining sample had a viral load of 1.62 log IU/mL. Two of the four high volume extracted samples in cohort 3 (with viral load 3.37 and 1.62 log_10_ IU/mL) were run on both the flongle R10.4 flowcell (FLO-FLG114) and the regular R10.4 flowcell, (FLO-MINI 114) to investigate reproducibility, and consensus sequences were identical from both flowcells.

**Table 2.** Analytical sensitivity and reproducibility after MagNA pure total NA extraction when running five pooled dilutions of one sample on a flongle flow cell.

| **Dilution** | **log<sub>10</sub> IU/mL** | **% of full genome** | **Similarity** | **Antiviral resistance detected\*** |
| --- | --- | --- | --- | --- |
| 1:1 | 5,34 | 100 | 100 | L180M, S202G, M204V |
| 1:10 | 4,34 | 100 | 100 | L180M, S202G, M204V |
| 1:100 | 3,34 | 56 | 100 | L180M, S202G, M204V |
| 1:1000 | 2,34 | 0 | NA | No |
| 1:10000 | 1,34 | 0 | NA | No |

\*Previous Sanger sequencing showed L180M, S202G and M204V.

### Sequence similarity

In cohort 1, the sequence similarity between Nanopore and Sanger was 99.9% (range 99.8-100%). Cohort 2 had not been Sanger sequenced. In cohort 3, the Sanger sequences covering the variable part of the RT were compared with the same regions in the Nanopore-derived sequences and showed a 99.8% similarity in mean (99.13-100%). Differences from Sanger were mainly caused by ambiguous bases in the Sanger-derived sequence.

### HBV splice variants and core deletions

Splicing at conventional splice sites was not detected in frequencies ≥10% threshold in any of the samples, however, reads suggesting splicing was found in 29 samples when the frequency threshold was lowered to 1%. Some of the HBV splice sites may affect the PCR target, most importantly the nt 2447-489 splicing junction, and could therefore potentially not be detected. Detection of all known preS and preCore deletions (26, 27) is however possible. Notably, in cohort 3, consisting of low viral load samples, one sample showed a preS1 deletion spaning nt 2847-3060, as the predominant genome variant, and one sample harboring an nt 2994-3054 deletion as a minor variant in approximately 25% of the reads (figure 2). No deletions were seen in cohorts 1 and 2.

**Figure 1.**
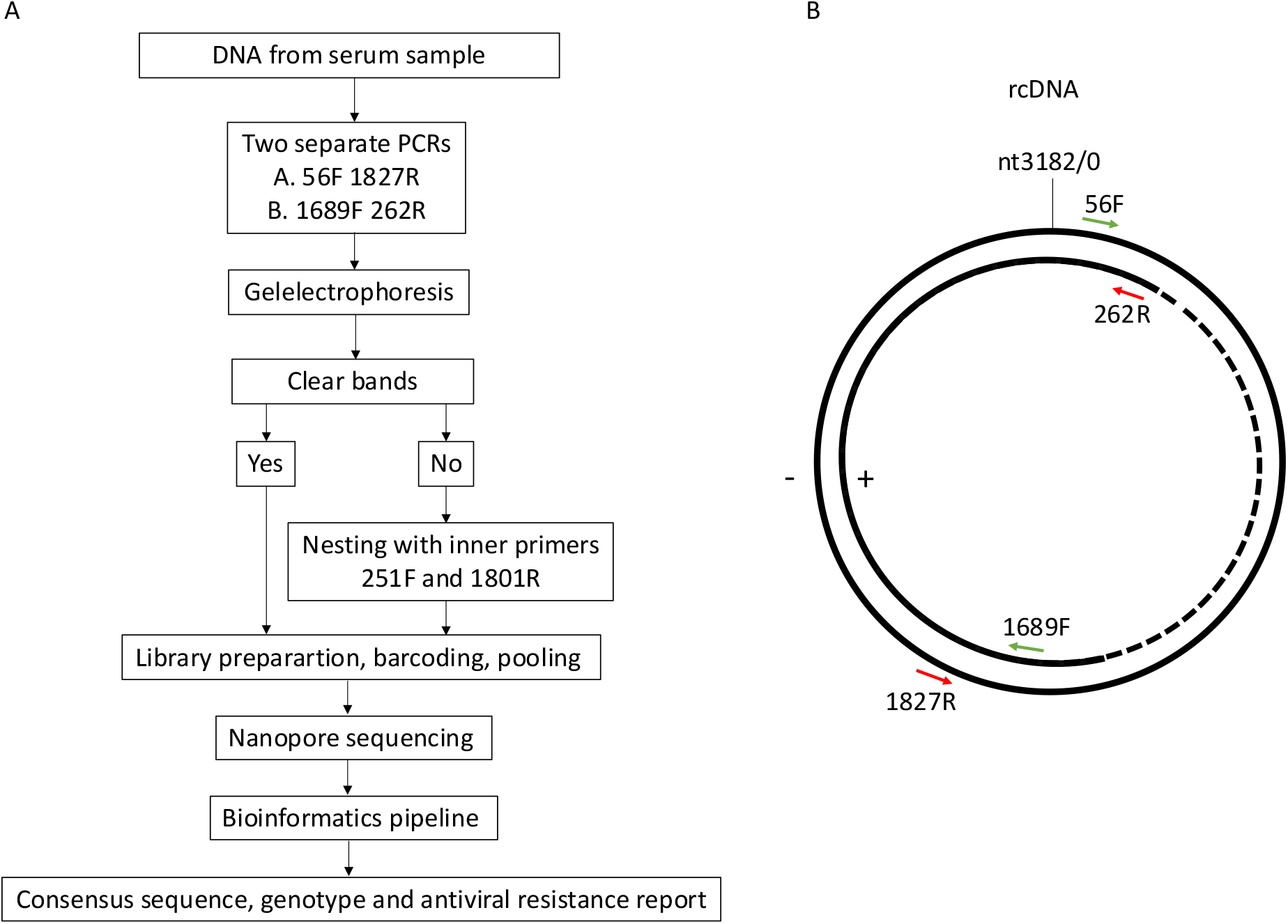
Method overview. A) Nanopore HBV whole-genome sequencing workflow. B) The position of the primers on the partially double-stranded HBV relaxed circular DNA (rcDNA), found in HBV particles. The method is designed to amplify both strands, including the shorter plus strand.

**Figure 2.**
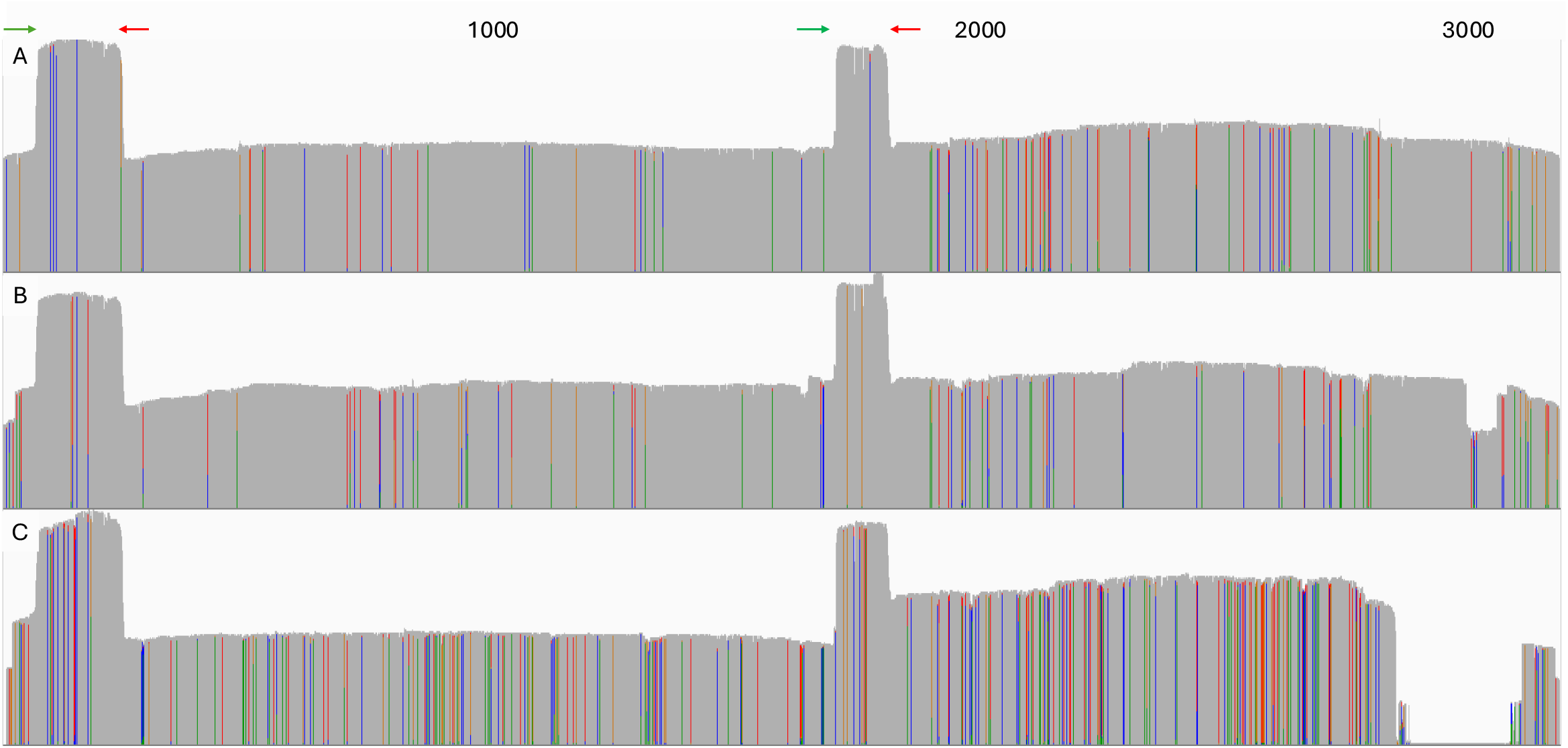
HBV coverage profiles from Nanopore sequencing of samples with and without structural aberrations. Colored lines indicate differences compared to the reference sequence reads were mapped against and grey matching nucleotides, the height reflects the number of reads covering the positions. The approximate primer locations are marked as green (forward) and red (reverse) arrows. A) Depicts a typical coverage profile, B) shows a sample with a preS1 deletion as a minor variant seen as a gap in a fraction of the reads at nt 2994-3054 and C) a sample with a majority of the HBV genomes with a preS1 deletion nt 2847-3060.

### Nanopore sequencing cost

Nanopore sequencing with the method described in this article can be performed with up to 96 samples per flow cell run or approximately twelve samples using the flongle flow cell. With current costs for conventional extraction and PCR (4€) and Nanopore reagents (R10.4.1 flow cell from 455€, flongle flow cell from 66€ and rapid barcoding kit cost per total barcoded library of 99€) the reagent cost for each run is in the range of 169-558 €, thus for each genome in the range of 6-37€, depending on flow cell used and the number of samples pooled.

## Discussion

Nanopore whole-genome sequencing of HBV was fast, sensitive and accurate, with 99.9% sequence agreement with Sanger sequencing. By introducing barcodes with the Nanopore rapid barcoding method, antiviral resistance, genotype and sequence analyses can be achieved at low cost. The main advantages of Nanopore are its simplicity and its feature to perform long-read sequencing. By amplifying and sequencing only two amplicons with lengths of approximately 1771 nt (nt 56 to 1827) and 1773 nt (nt 1689 to 262), respectively, the whole circular HBV genome was captured. The primer regions were chosen to obtain continuous reads for the RT, precore-core region and for the regions potentially involved in vaccine escape and antiviral resistance (28–30).

For this study, we used the Rapid Barcoding kit 96 V14 but alternative Nanopore kits are also possible. The ligation sequencing-based kits were tested during the method development but tend to be more laborious and time consuming, however, the amplicons remain intact and are sequenced as one continuous read, which could be preferable in haplotype and structural variation analysis. Despite this transposase effect, we believe that the rapid barcoding kit is best suited for routine diagnostics, as it demands little hands-on time and may be used for sequencing and pooling of several different microbes simultaneously, which may be either PCR-enriched or cultured. The same chemistry has also been used for SARS-CoV-2 sequencing and has proven to be scalable to up to 96 samples in one run (31).

We tested two different nucleic acid extraction protocols in this study, and the modified protocol for the manual high-volume kit from 5 mL of plasma by Fu *et al*. (23) proved to be useful in low viral load samples, and full genome coverage was achieved from samples as low as 1.62 log10 IU/mL, that could not be achieved with MagNA pure 96 extraction from 200 microliters of serum. Presumably, the high-volume kit yielded higher levels of intact DNA and, therefore, was preferable for long-read sequencing. Serial dilution of one sample showed that for concentrations ≥3.43 log10 IU/mL, Nanopore sequencing of MagNA pure 96 (Roche) extracts showed reproducible results and allowed detection of resistance mutations. However, sequencing of the whole RT gene was achieved in all samples of cohort 1 and in contrary to the serial dilutions, full genomes were captured in eight samples with viral load <3.43 log10 IU/mL also after MagNA pure 96 extraction. The seemingly lower sensitivity in the serial dilution run compared to the other Nanopore runs might be explained by the differences in viral concentrations, causing amplicons from higher concentration dilutions to outcompete those with lower in the pooled reaction. The sensitivity could possibly be increased if samples with high concentrations are diluted, but this did not seem necessary in the other samples and runs for which sufficient results were obtained.

We applied the GPU-based super accurate basecalling for fast yet high-quality generation of reads. Nanopore basecalling has improved immensely in recent years and is the main factor for lowered error rate and several alternative methods have been presented (32–35). In a previous study, we used a pipeline for consensus sequence extraction from NGS data and almost perfect sequences were achieved, but some false insertion/deletions (indels) remained (3). To overcome this issue in more error prone long-reads data, the Oxford Nanopore Technologies tool Medaka was used to create a consensus sequence from Minmap2-mapped. The implementation of the Medaka tool removed false indels, as previously shown for SARS-CoV-2 (19). Medaka creates consensus sequences based on the major virus variant and for minor variant analysis, we applied freebayes (24). In our analysis of antiviral resistance all variants that previously had been found using Sanger sequencing were found and no false positives were detected applying a frequency threshold of 10%. None of the samples in the study had resistance mutations constituting <25% of the viral population, supporting the threshold used. Previous studies of Nanopore error-rate suggest that this could be lowered to 3% (36), something we aim to investigate in future studies.

Previous studies have presented sequencing of HBV combined with rolling-circle amplification (37, 38). McNaughton *et al*. adapted the method for Nanopore sequencing and minor variant analysis (38). Their approach with rolling-circle amplification resulted in extremely long amplicons and very long reads with the whole HBV sequence repeated, optimal for haplotype reconstruction and high accuracy. For routine diagnostics a standard PCR approach might still be preferable, as it presumably yields higher sensitivity (37) and less laborious protocols (38). The method presented in this article, based on conventional PCR, had an almost 100% agreement between Nanopore and Sanger derived consensus genomes, allowing for clinical use. If detection of ultra-rare minor variants, or *in haplo* mutations, is needed for research purposes, we believe that the method presented by McNaughton *et al*, Ion Torrent or Illumina-based methods, might be preferable (39).

This study showed that deletions in HBV genomes can be found using long-read sequencing. Deletions have been shown in many studies but their clinical relevance is not fully known (40). Also, a recent article suggests that splicing is associated with poorer interferon treatment outcomes (41). HBV genome deletions, such as those we found in the S region in two samples in this study, have been associated with an increased risk of cirrhosis (42). We present a method that enables large-scale use and characterization of structural genomic features, which allows for bigger studies. Other primers than those presented here could improve the detection of splicing variants (42–44) and we plan to study this.

Other limitations of this study include the lack of Sanger sequencing covering the entire HBV genome meaning only RT region could be compared with Nanopore. Also, the primers for the second amplicon in the Nanopore method, covering the S region, are not optimal for genotypes F-J as some strains have up to three mismatches in the primer target, discovered after the completion of this study. However, the primers for the RT amplicon appear to be conserved in all genotypes which enable pan-genotypic resistance and genotyping. The aim of this article was to present a method suitable routine laboratories and thus only used a nested PCR for the amplicon covering the RT region, however, the whole genome sensitivity may be further increased for all genotypes if additional amplicons are sequenced (45) or nesting of the second amplicon is applied.

Our study shows that Nanopore sequencing of two overlapping amplicons is a feasible method for HBV whole-genome sequencing, genotyping and antiviral resistance mutation analysis in a routine laboratory setting, showing adequate sensitivity and accuracy for clinical purposes. Minor variant analysis of clinically relevant subpopulations was possible. We believe that this method, providing simple and cost-efficient sequencing, may contribute to increased whole-genome sequencing of HBV and new discoveries of clinically relevant genomic features.

## Data Availability

All data produced in the present study are available upon reasonable request to the authors.

## Author contributions

JR, HH and UN planned and designed the study. JR, MA and JBS designed the Nanopore method. JR and PN financed the study. JBS, UN, MA and MF performed laboratory work. JG designed hardware and set up GPU-based basecalling. DS, JR and JBS designed and performed bioinformatical analyses. JR drafted the manuscript. All authors contributed to the final version of the manuscript.

## Data availability

The data that support the findings of this study are openly available at the European Nucleotide Archive (ENA) https://www.ebi.ac.uk/ena/browser/home. Fastq files are uploaded with the study accession number PRJEB87509.

